# A Review of Dosages of Chloroquine and Hydroxychloroquine for COVID-19 in registered Clinical Trials during First Quarter of 2020

**DOI:** 10.1101/2020.03.22.20040964

**Authors:** Divya RSJB Rana, Santosh Dulal

## Abstract

**Background:** The novel corona virus disease 2019 (COVID-19) pandemic has been causing a massive global public health havoc. Use of quinolones for treatment of COVID-19 was a matter of huge discussion in scientific community. Falsified data about efficacy of the drug against COVID-19 disseminated. This review was designed to study the dosages of chloroquine and hydroxychloroquine planned to be administered in clinical trials registered up to March 2020.

**Summary:** Inclusion of chloroquine and hydroxychloroquine for COVID-19 treatment in Chinese national treatment guideline in the early days of the pandemic prompted numerous clinical trials in many countries to authenticate the efficacy of the drugs. Trials were designed to include chloroquine or hydroxychloroquine singly or in combination with other drugs. Almost all of the trials planned oral administration except few which used aerosol inhalation. In the later half of 2020, systematic reviews and results of those clinical trials point out the inefficacies and inadvertent adverse events due to the use of these quinolone drugs for COVID-19.

**Conclusion:** This study reviews the various dosages of chloroquine and hydroxychloroquine utilized in published and under-study clinical trials as assessed during the end of March 2020. Specifically, clinical trials registered in Chinese and US trial registries were examined.

## Background

Coronavirus disease 2019 (COVID-19) is an infectious disease of respiratory tract of potential zoonotic origin ^1^□. Severe acute respiratory syndrome (SARS) corona virus – 2, (SARS-CoV-2), the etiological agent of COVID-19, causes a serious or life-threatening disease or condition, including severe respiratory illness ^1^□□□. The public health concerns caused by the outbreak of COVID-19 was declared as a “public health emergency of international concern” on 31 January and “pandemic on 11 March 2020 by World Health Organization ^2^□. Impacts of the COVID-19 pandemic touched all the corners of the globe leading to colossal economic and public health crisis. The reported mortality rate for COVID-19 in the early days was around 3% ^3^□ and the mortality rates vary depending on geographical regions, age distributions and co-morbidities ^4^□ making it a subject of debate ^5^□□.

In the early days of the pandemic, the Chinese Government included chloroquine phosphate in the list of antiviral therapies for the treatment of novel coronavirus pneumonia or COVID-19 caused by severe acute respiratory syndrome coronavirus 2 (SARS-CoV-2) in its 6th and 7^th^ versions of the official guidelines for the Prevention, Diagnosis, and Treatment of Pneumonia caused by COVID-19 ^6,7^□□□.

Conclusive research on the potential therapeutic applications for the disease could have saved lives and prevent the rapid viral transmission. Chloroquine phosphate is an old drug used for the treatment of malaria caused by *Plasmodium species* ^*8*^□□. Chloroquine phosphate can also be replaced with another member of its family, hydroxychloroquine ^9^□. The active compound chloroquine constitutes 60% in chloroquine phosphate and 77.5% in hydroxychloroquine respectively.

Earlier in vitro studies had shown efficacy of chloroquines against coronaviruses. Mechanism of actions of chloroquine or hydroxychloroquine had been studied for both SARS-CoV and SARS-CoV-2. A study using Vero E6 cells found that treatment of the cells with chloroquine either before or after infection with SARS-CoV could prevent infection or treat the infected cells suggesting both prophylactic and therapeutic effect ^10^□□□. The study found chloroquine disturbed the biochemical maturation of the cellular receptor, angiotensin converting enzyme 2 (ACE2), for SARS-CoV, and increased the pH of intracellular virus-laden vesicles inhibiting the viral normal multiplication processes. The receptor for SARS-CoV-2 is also highly predicted of being ACE2 □ ^1^□. In vitro studies for SARS-CoV-2 also found chloroquine and hydroxychloroquine, a less-toxic derivative of chloroquine to be able to effectively inhibit viral growth ^11^□. They found the viral biological processes in the endosomes were affected by the anti-malarial agents. While the reported 50% effective concentration of the drugs were higher than the normal serum concentration of drugs in people taking these drugs, the concentrations in lungs and other vital organs are higher ^12^□ and could lend support for the idea that the drug could perhaps be useful□. The most common side-effect of these are retinopathy and other side-effects precipitating due to higher concentrations of the compounds in blood.

The recommended 3-day chloroquine phosphate dosage for malaria treatments for adults is 1 gm orally followed by 500 mg after 6-8 hours in the first day and 500 mg per day for second and the third day ^8^□. Similarly, the 3-day dosage for hydroxychloroquine is 800 mg orally followed by 400 mg after 6-8 hours in the first day and 400 mg per day for second and third day ^9^□.

This study was specifically conducted to examine the various dosages of chloroquine and hydroxychloroquine utilized in clinical trials registered in China and USA for the treatment of pneumonia caused by SARS-CoV-2. The guidelines by the Chinese government has postulated the dose of 500 mg twice a day for 7 days for adults aged 18-65 years and weighing more than 50 kg; and initial dose of 500 mg twice a day for first two days and 500 mg per day for days 3 through 7 for adults of weight less than 50 kg ^7^□.

## Methods

A systematic search of registered clinical trials was carried (Figure 1, PRISMA flow-chart). The list of clinical trials registered in China in a ready-made excel spreadsheet format was downloaded from the Chinese clinical trial registry ^13^□ on 30^th^ March, 2020 provided in the website. The list of clinical trials registered in the US government clinical trial registry ^14^□□ were retrieved by searching “coronavirus” as keyword on 30^th^ March, 2020. All the trials were filtered out based on the presence of word “chloroquine” in the title irrespective of the chemical forms of the molecule (hydroxychloroquine) or trials being carried alone or in combination with other antiviral therapies. An excel spreadsheet file was prepared based on various headings including intervention groups, randomization status, sample sizes, etc. Univariate statistics was used to report the methodologies of the trials.

**Figure 1:**
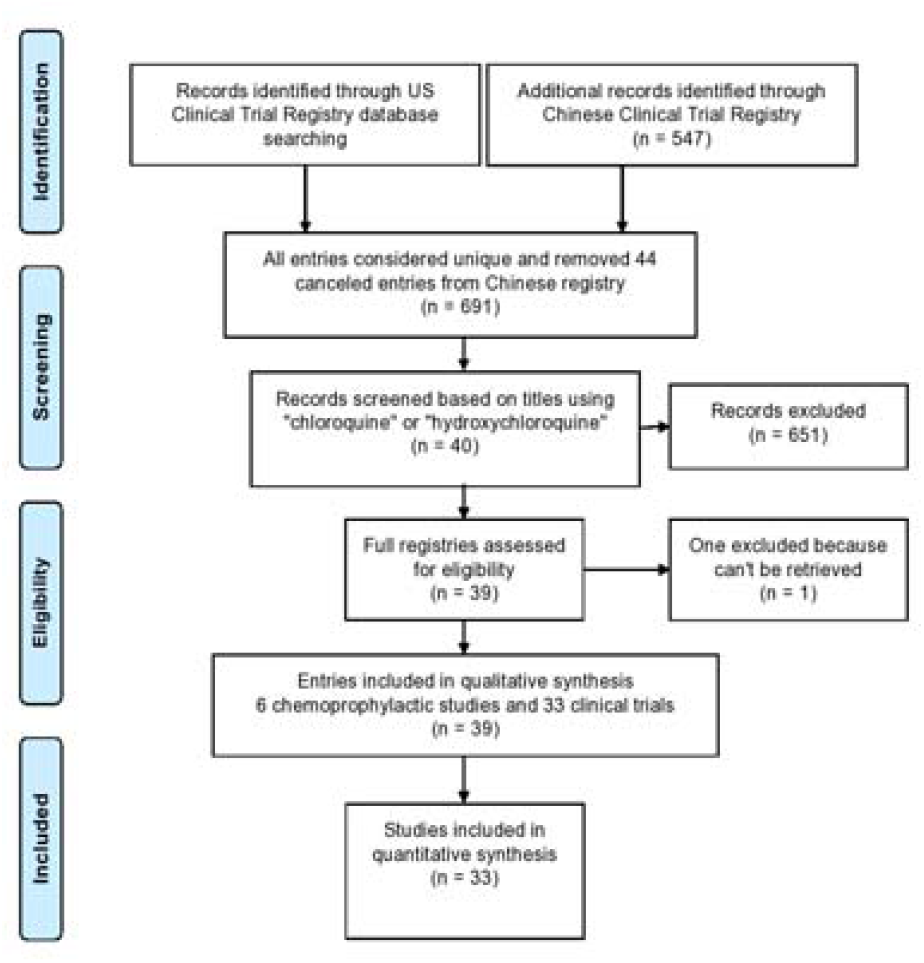
PRISMA flow diagram for strategy for search of registered clinical trials

## Results

As of 30^th^ March 2020, 547 trials were registered in the Chinese clinical trial registry starting from 23^rd^ January, 2020. 25 (4.6%, 25/547) clinical trials used the word chloroquine in their titles. Five of them were reported to had been canceled, one was a prophylactic study and one another trial information could not be retrieved. Eighteen (3.3%, 18/547) clinical trials from the Chinese registry were used for the downstream analysis. As of 30^th^ March 2020, 184 entries were obtained by searching for coronavirus related clinical trials in the US clinical trial registry of which only 153 were registered after 1^st^ January 2020. Out of 153 registrations, 20 (13.1%, 20/153) were found to use chloroquine. Four of these were registered from China, 3 from Brazil, 2 each from France, Mexico, US and Canada, and 1 each from Thailand, South Korea, United Kingdom, Norway and Turkey. Two of the studies from US, and 1 each from the United Kingdom, Mexico and Turkey were prophylactic study for healthy people at risk of infection. Thus 15 studies from US trial registry were used for analysis. Total 33 (18+15) clinical trials utilizing the quinolones had been registered since 3rd February 2020. Six prophylactic studies are analyzed separately.

Most of the trials proposed oral chloroquine (68.2%, 15/33) or hydroxychloroquine (68.2%, 15/33) singly as treatment options, followed by chloroquine in combination with other antivirals ((13.6%, 3/33), hydroxychloroquine with azithromycin (9.1%, 2/33), chloroquine as aerosols (9.1%, 2/33), chloroquine with azithromycin (4.5%, 1/33) and hydroxychloroquine with other antivirals (4.5%, 1/33) (Table 1). The maximum quantity of chloroquine planned was 12 g in a period of 10 days (NCT04323527) and the lowest reported was 2.1 g aerosolized chloroquine (ChiCTR2000029975) in a duration on 7 days. Similarly, the maximum quantity of hydroxychloroquine proposed was 12.4 gm in a duration of 14 days (ChiCTR2000029868) and the lowest amount was 2 gm in a period of 5 days (NCT04261517). Only one of the two trials proposing aerosolized chloroquine phosphate stated the dosage. It was 2.1 gm in a period of 7 days (ChiCTR2000029975). A total of 12,894 study patients were planned to be enrolled in 33 trials. Twelve trials registered in the US registry didn’t have details about sample size allocation into individual intervention arms. For the 5,842 individuals in 21 trials with information on sample sizes in individual intervention arms, 1199 patients were to get chloroquine alone, 1050 were to get hydroxychloroquine alone, 159 were to get chloroquine with other drugs, 25 were to get aerosolized chloroquines, 1320 were to get conventional treatments or placebo, and 2089 were to get other antivirals. All of the active or placebo interventions were planned to get conventional treatments or the standard care. Out of the 33 clinical trials, 4 reported that they were not designed to be randomized, two were single arm studies and didn’t require randomization and 27 reported using some kind of randomization. Out of total, nine-tenth (91%, 30/33) of the studies had set 18 years old as lower age limit for inclusion criteria while two trials used 16 years, and one trial used 30 years. Similarly, upper age limit ranged from 65 to 100 years of age. All trials had planned to enroll both genders in the study.

**Table 1:** Use of different forms of chloroquine in clinical trials

| Type of trials | Trials (%)* |
| --- | --- |
| Chloroquine phosphate | 15 (68.2%) |
| Chloroquine phosphate with other antivirals | 3 (13.6%) |
| Chloroquine with azithromycin | 1 (4.5%) |
| Hydroxychloroquine | 15 (68.2%) |
| Hydroxychloroquine with others | 1 (4.5%) |
| Hydroxychloroquine with azithromycin | 2 (9.1%) |
| Chloroquine phosphate as aerosol inhalation | 2 (9.1%) |
| Total | 33 (100%) |

\*Few trials have one or more types together in same study

**Table 2:**
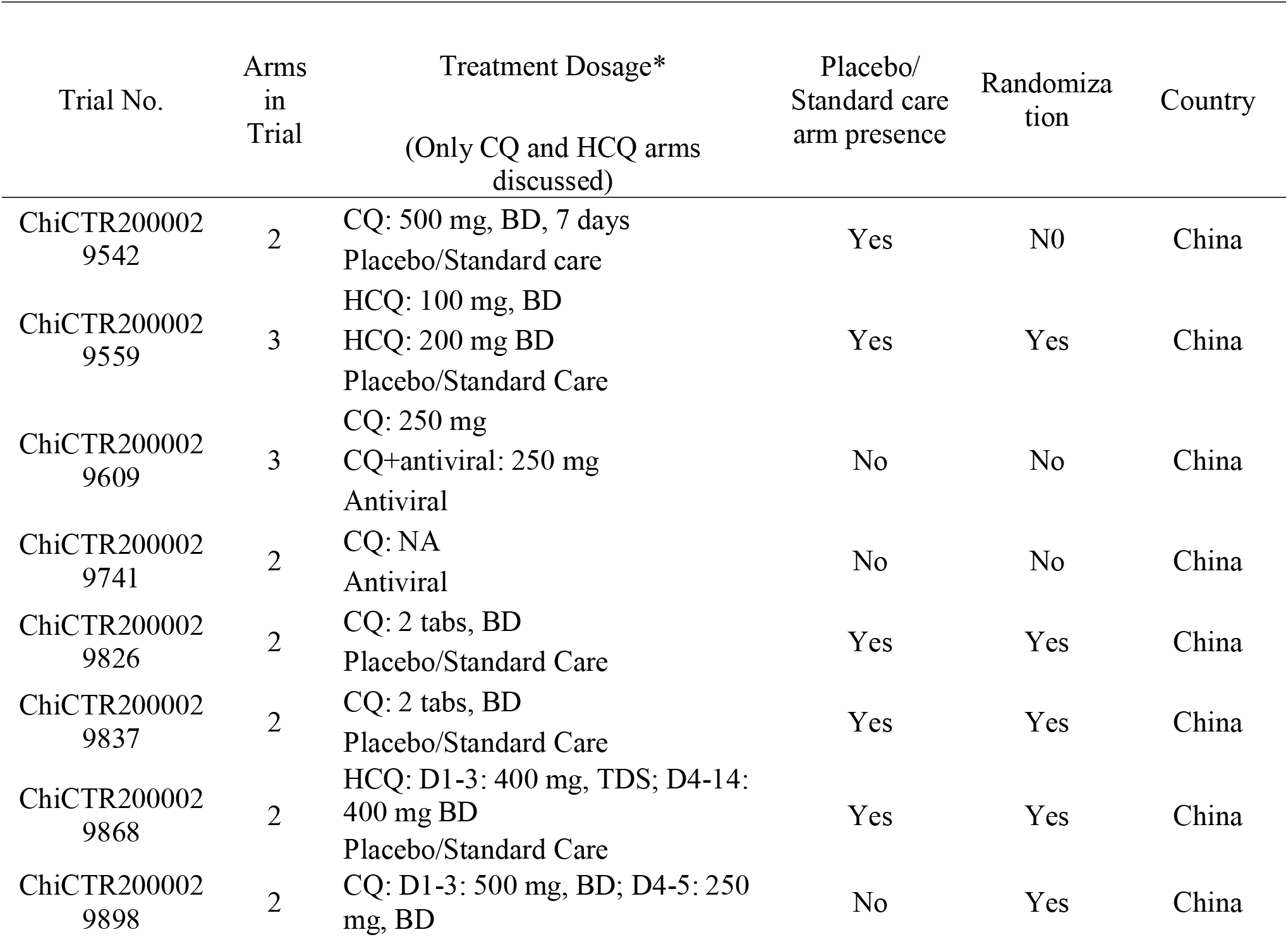
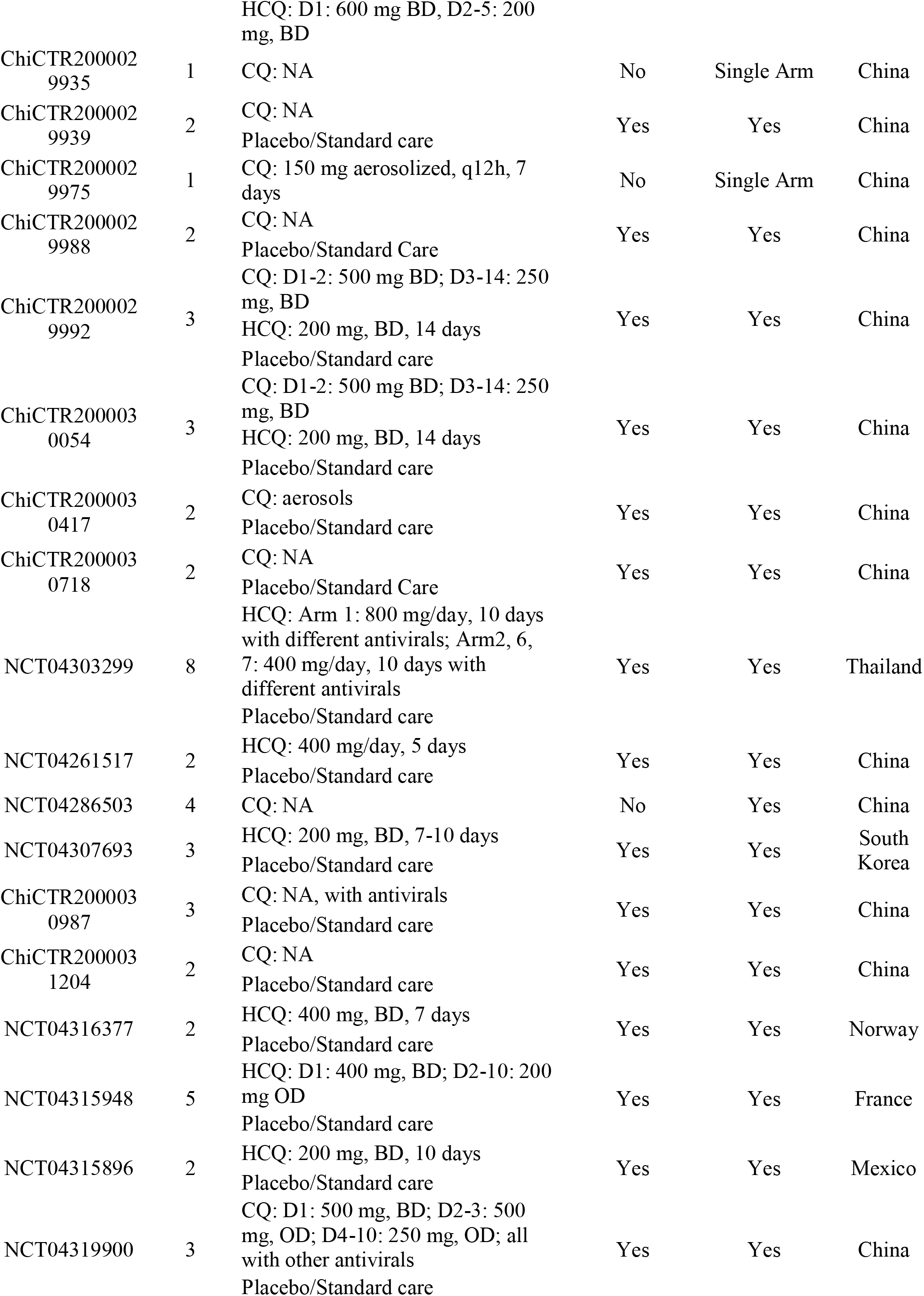
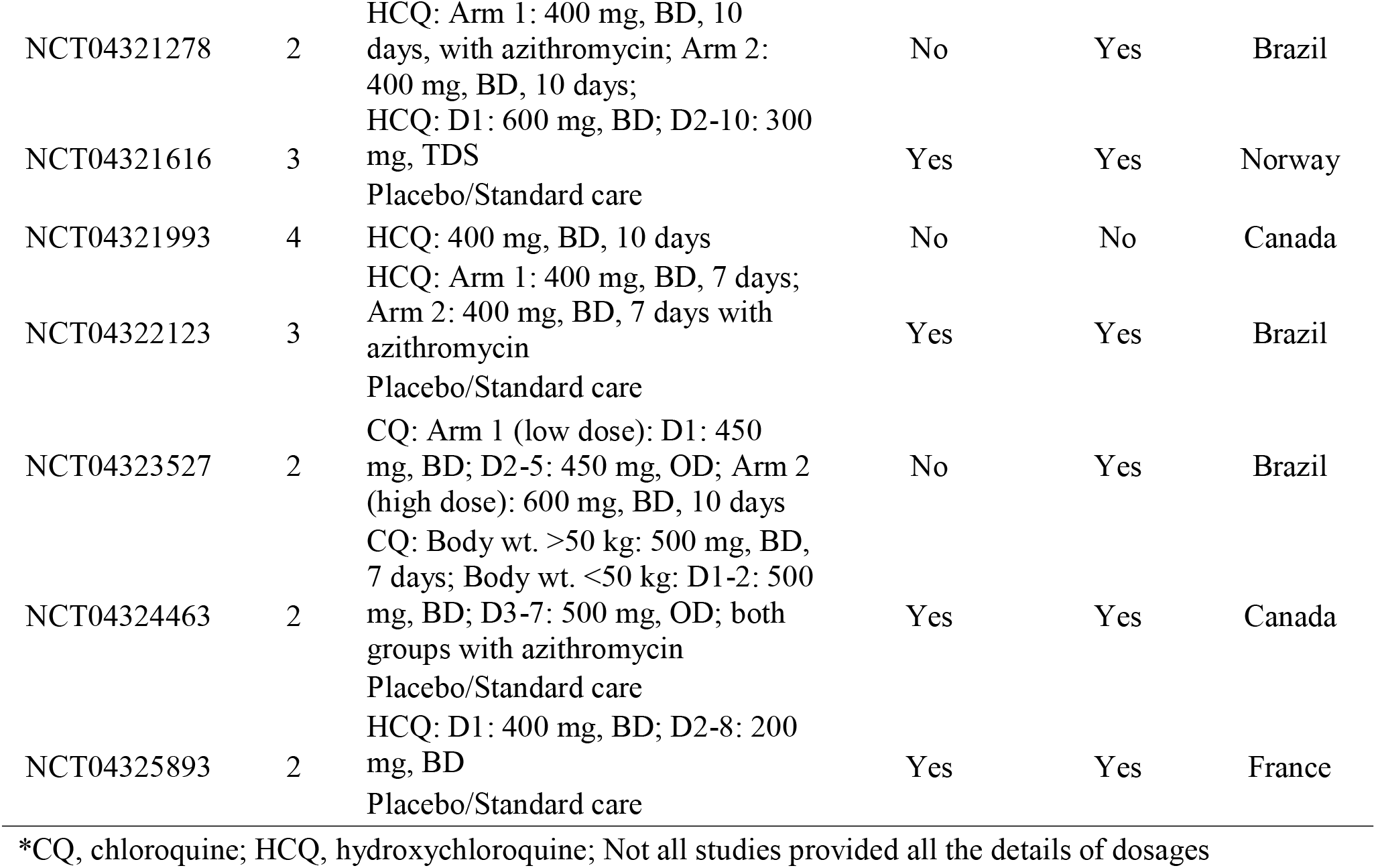
Dosages and characteristics of the registered clinical trials

Among the trials using chloroquine or hydroxychloroquine as prophylactic agent (Table 3), 45,400 healthy at-risk population were to be enrolled, among which the UK study by the University of Oxford, planned to enroll 40,00 individuals for a duration of three months with 250 mg chloroquine daily (NCT04303507). A small dose of 200 mg hydroxychloroquine given every three weeks along with daily vitamin C and zinc tablets) (NCT04326725) was planned in an another prophylactic study in Turkey. The ambitious projects hoped to provide low-cost option for vaccination which could not save millions of lives in coming months.

**Table 3:**
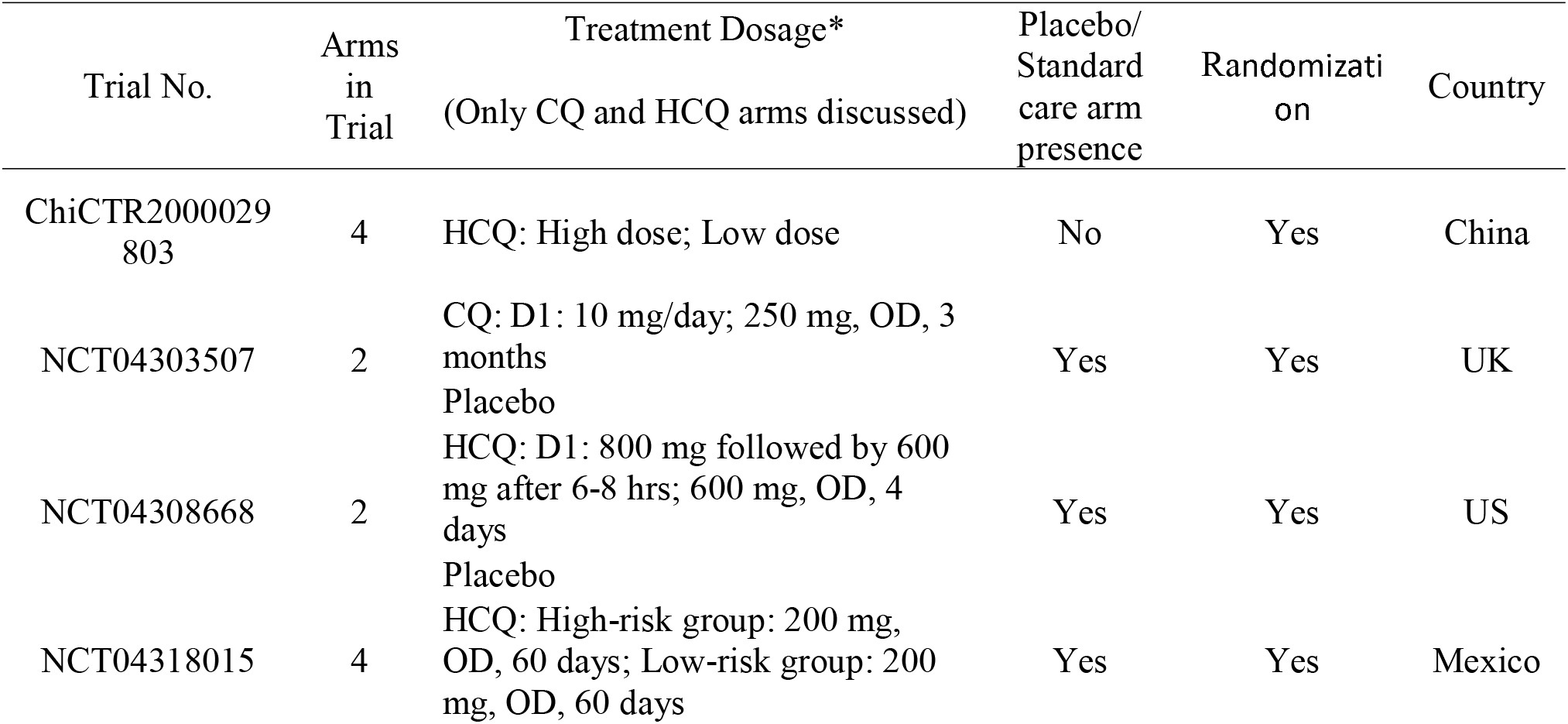
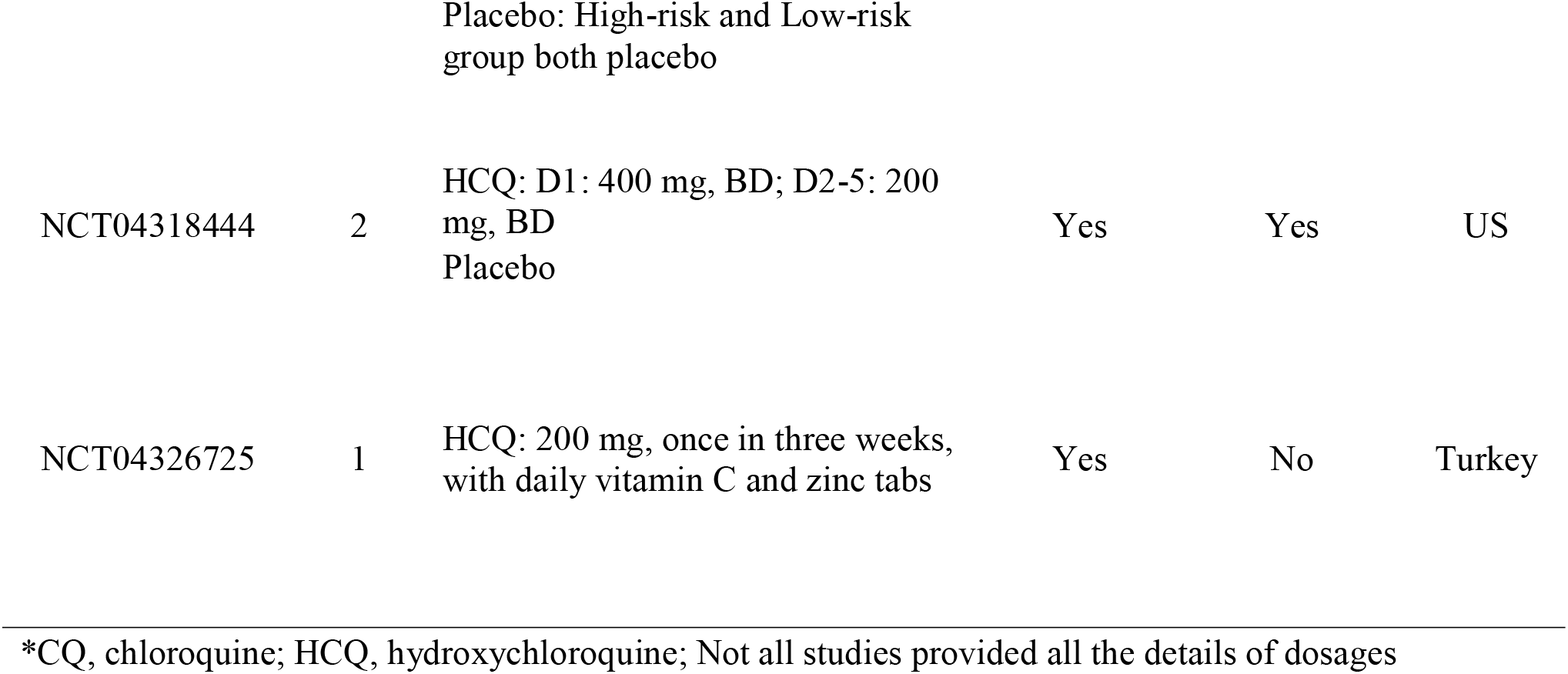
Dosages and characteristics of prophylactic clinical trials

The background information in a Chinese clinical trial (ChiCTR2000029741) ^15^□ had revealed that choloroquine phosphate (5 days medications) was fifty percent (5/10) effective compared to 20% (3/15) for combination medication of lopinavir/ritonavir (5 days medications) in Chinese COVID-19 pneumonia patients in 2020.

The Chinese government in its 6^th^ and 7^th^ guidelines for the treatment of novel coronavirus pneumonia had recommended use of chloroquine phosphate at specific dosages coinciding with a very brief summary article from China ^16^□ about successful treatment of COVID-19 patients with chloroquine in China. One of the first trials using chloroquine as treatment drug for COVID-19 outside China was a non-randomized open-label clinical trial in France that enrolled hospitalized COVID-19 patients to the hydroxychloroquine (20 patients) and control arms (16 patients) concluded a positive effect of hydroxychloroquine or hydroxychloroquine plus azithromycin based on viral negativity at day 6 ^17^□□.

The study played role in prompting the US FDA to give emergency use authorization for the drug ^18^□. The study was later hugely criticized due to inappropriate patient enrollment, study design, and unethical conduct of an editor of the journal being one of the author of the article. The scientific society related to the journal shared the concern ^19^□. A review noted the inclusion of chloroquine/hydroxychloroquine based therapy in the national guidelines of China, Korea, Netherlands, Italy, Canada and Belgium ^20^□□. RECOVERY trial by the University of Oxford in the UK which enrolled 11 thousand hospitalized COVID-19 patients concluded no benefit of hydroxychloroquine ^21^□. A systematic review and meta-analysis of seven randomized control trial concluded no benefit of hydroxychloroquine for mild and moderate forms of COVID-19 disease and increase in adverse events in the treatment group ^22^□. A Cochrane review of clinical trials found no benefits of using the quinolones on mortality and mechanical ventilation of the COVID-19 patients ^23^□. Moreover the review found increased proportion of adverse events in the treatment arm.

## Conclusion

Currently, there are very limited information and knowledge regarding the therapeutic aspects of COVID-19 as most of the clinical trials are still undergoing and their findings have not been publicly available yet. Touret and de Lamballerie ^24^□ reported the scarcity of data in clinical trials on therapeutic aspects of chloroquine or hydroxychloroquine. They pointed out that in spite of wonderful ex vivo activities of the quinolones against influenza, dengue, chikugunya, etc. the effect failed to replicate in in vivo studies. Gao et al. ^16^□ □in an abstract-like article revealed that chloroquine phosphate had superior efficacy for controlling the inhibition of pneumonia exacerbation, disease progression and transmission in more than 100 Chinese COVID-19 patients. Scientifically sound studies and results of time-tested clinical trials are required for conclusive identification of treatment regimens. Recent and previous *in vitro* studies have shown the effectiveness of chloroquine and hydroxychloroquine against COVID-19 or other coronaviruses (Table 4) and these data prove that in results of vitro studies may not be always replicated in clinical trials.

**Table 4:** In vitro studies using chloroquine/hydroxychloroquine against coronavirus.

| Virus | Host | Drug | EC50* (mcM) | Author, Year |
| --- | --- | --- | --- | --- |
| SARS-CoV-2 | Vero | Hydroxychloroquine sulf. | 0.72 (poi) | Yao 2020 |
| SARS-CoV-2 | Vero | Chloroquine phosp. | 5.47 (poi) | Yao 2020 |
| SARS | Vero E6 | Chloroquine | 4.1 (poi) | de Wilde 2014 |
| MERS | Huh7 | Chloroquine | 3.0 (poi) | de Wilde 2014 |
| HCoV-229E | Huh7 | Chloroquine | 3.3 (poi) | de Wilde 2014 |
| SARS | Vero E6 | Chloroquine | 1 mcM (pri)_ | Vincent 2005 |
| SARS | Vero E6 | Chloroquine | 4.4 mcM (poi) | Vincent 2005 |
| rHCoV-OC43 | BHK-21 | Chloroquine | IC50 0.33 (poi) | Shen 2016 |
| HCoV-OC43 | HRT-18 | Chloroquine phosp. | 0.306 mcM (poi) | Keyaerts 2009 |
| MERS | Dendritic cell | Chloroquine diphosp. | Not effective | Cong 2018 |
| SARS | Vero E6 | Chloroquine | IC50 8.8 (poi) | Keyaerts 2004 |
| FIPV | CRFK | Chloroquine diphosp. | IC50 16.63 (pri) | McDonagh 2014 |
| FIPV | CRFK | Chloroquine diphosp. | IC50 0.38 (24h pri) | McDonagh 2014 |
| FIPV | CRFK | Chloroquine diphosp. | IC50 28.87 (48h pri) | McDonagh 2014 |
| MERS | Vero E6 | Chloroquine diphosp. | 6.3 (pri) | Dyall 2014 |
| SARS | Vero E6 | Chloroquine diphosp. | 6.5 (pri) | Dyall 2014 |
| MERS | Vero E6 | Hydroxychloroquine<br>sulf. | 8.3 (pri) | Dyall 2014 |
| SARS | Vero E6 | Hydroxychloroquine<br>sulf. | 7.9 (pri) | Dyall 2014 |
| SARS | Vero 76 | Chloroquine | 2.5 (sti) | Barnard 2006 |
| SARS | Vero 76 | Chloroquine diphosp. | 6.0 (sti) | Barnard 2006 |
| SARS-CoV-2 | Vero E6 | Chloroquine | 1.13 (pri) | Wang 2020 |

50% effective concentration unless stated; IC50: 50% inhibitory concentration; poi: post infection; pri: pre infection; sti: drug and infection at same time
\*\*References in supplementary information

Continued efforts to carry ethically valid clinical trials are required for the assessment of effective and safe dosage administration. This can importantly contribute to guide the medical professionals for prescribing the accurate interventions and also contribute to pharmaceutical companies for deciding appropriate manufacturing pathways.

## Data Availability

No extra data required.

## Contributions

SD and DR conceptualized the study. SD laid out the plan of the study. DR wrote the first draft. SD and DR edited and agreed on the final draft.

